# Memory recovery is related to default mode network impairment and neurite density during brain tumours treatment

**DOI:** 10.1101/19008581

**Authors:** Rafael Romero-Garcia, John Suckling, Mallory Owen, Moataz Assem, Rohitashwa Sinha, Pedro Coelho, Emma Woodberry, Stephen J Price, Amos Burke, Thomas Santarius, Yaara Erez, Michael Hart

## Abstract

**Objective:** The aim of this study is to test brain tumour interactions with brain networks thereby identifying protective features and risk factors for memory recovery after surgical resection.

**Methods:** Seventeen patients with diffuse non-enhancing glioma (aged 22-56 years) were longitudinally MRI-scanned before and after surgery, and during a 12-months recovery period (47 MRI in total after exclusion). After each scanning session, a battery of memory tests was performed using a tablet-based screening tool, including free verbal memory, overall verbal memory, episodic memory, orientation, forward digit span and backwards digit span. Using structural MRI and Neurite Orientation Dispersion and Density Imaging (NODDI) derived from diffusion-weighted images, we respectively estimated lesion overlap and Neurite Density with brain networks derived from normative data in healthy participants (somato-motor, dorsal attention, ventral attention, fronto-parietal and Default Mode Network -DMN-). Linear Mixed Models (LMMs) that regressed out the effect of age, gender, tumour grade, type of treatment, total lesion volume and total neurite density were used to test the potential longitudinal associations between imaging markers and memory recovery.

**Results:** Memory recovery was not significantly associated with tumour location based on traditional lobe classification nor with the type of treatment received by patients (*i*.*e*. surgery alone or surgery with adjuvant chemoradiotherapy). Non-local effects of tumours were evident on Neurite Density, which was reduced not only within the tumour, but also beyond the tumour boundary. In contrast, high preoperative Neurite Density outside the tumour, but within the DMN, was associated with better memory recovery (LMM, *P*_*fdr*_<10^−3^). Furthermore, postoperative and follow-up Neurite Density within the DMN and fronto-parietal network were also associated with memory recovery (LMM, *P_fdr_*=0.014 and *P_fdr_*=0.001, respectively). Preoperative tumour, and post-operative lesion, overlap with the DMN showed a significant negative association with memory recovery (LMM, *P_fdr_*=0.002 and *P_fdr_*<10^−4^, respectively).

**Conclusion:** Imaging biomarkers of cognitive recovery and decline can be identified using NODDI and resting-state networks. Brain tumours and their corresponding treatment affecting brain networks that are fundamental for memory functioning such as the DMN can have a major impact on patient’s memory recovery.

## INTRODUCTION

Every year more than 300,000 people worldwide face the diagnosis of a brain tumour. Patients who undergo surgical resection rather than biopsy have a better overall survival, and extending the resection beyond the abnormality seen on MRI may further improve prognosis. However, the extent of resection is only a worthwhile prognostic factor in the management of the tumour if subsequent cognitive functioning can be maintained. Cognitive function, despite being recognised as a fundamental outcome measure of treatment success, remains one of the most unpredictable aspects of patients’ prognosis. As a result of their tumours, a significant proportion of patients develop cognitive deficits ranging from 29% in patients with non-irradiated low-grade glioma, to about 90% in patients following chemo-radiotherapy for metastases^1^. Among the wide range of acquired cognitive deficits, memory disabilities represent a major challenge for preserving patients’ quality of life, including job preservation^2^. Working^3^, verbal^4^ and non-verbal^5^ memory deficits have been extensively reported in brain tumour patients. Pharmacological interventions improve memory recovery^6^, which has been validated in animal models^7^. Unfortunately, the impact of surgery on cognitive outcome and memory has traditionally been underappreciated, potentially due to the complex distributed networks involved in normal memory function, and the limited understanding on how focal lesions interact with these networks^8^.

Typical MRI sequences used for clinical evaluation are limited by low biological specificity that constrains their capability to differentiate tumour types and infiltration as well as determine macromolecular and histological compositions. For example, although gadolinium contrast enhancement is currently used with MRI as a marker of anaplastic transformation of diffuse lower-grade gliomas, up to one-third of high-grade gliomas are non-enhancing^9^. Alternatively, diffusion MRI sequences probe the random (Brownian) motion of extracellular water and the restricted diffusion of intracellular water to determine brain tissue microstructure and orientation. Diffusion MRI protocols have been exploited for estimating tumour infiltration, grading, heterogeneity, progression and patient survival^10^, suggesting that the incorporation of these MRI protocols into the clinical routine may also be useful in reducing treatment-induced neurocognitive dysfunction in brain tumours. Notwithstanding, most previous brain tumour studies have used diffusion models under the assumption of Gaussian diffusion processes. In contrast, the Neurite Orientation Dispersion and Density Imaging (NODDI) technique uses varying diffusion gradients strengths to provide more specific measurements of neurite morphology such as density and orientation dispersion that cannot be derived from traditional diffusion MRI^11^. NODDI models the diffusion signal by separating the brain tissue into three compartments: intraneurite water, extraneurite water and cerebrospinal fluid. The volume fraction of the intraneurite compartment has been established as a marker of Neurite Density that has validated using histochemical analysis in mouse models^12^ and has shown sensitivity to neurotypical ageing^13^ and for detecting axonal degeneration in neurological (such as Alzheimer’s^14^, multiple sclerosis^15^) and psychiatric conditions^16^. Caverzasi *et al*. (2016) showed that even for tumour lesions that appear to be homogeneous on corresponding fluid-attenuated inversion-recovery (FLAIR) images, NODDI has the potential to differentiate infiltrative tumour components^17^. Additionally, NODDI has been demonstrated to be an accurate predictor of glioma grade^18^. Despite these promising findings, no study has yet used NODDI to evaluate the impact of tumours and their treatment on cognition, including memory.

A major challenge when predicting the consequences of surgery and chemo-radiotherapy is that higher-order cognitive functions such as memory are sub-served by communication across spatially extended neural circuits that will require whole-brain analysis approaches. Large-scale brain networks offer a holistic framework to analysing the brain as a circuit of interacting components that are critically related to cognition^19^. In neuro-oncology, markers derived from brain network approaches have been demonstrated to be sensitive to the presence of low-grade gliomas, plasticity differences between low- and high-grade gliomas, and surgically induced alterations^20^. Consequently, the use of brain network data to explore the potential for memory disruption induced by brain tumours and their treatment could be of major clinical relevance.

In this longitudinal study, we prospectively investigated a cohort of patients with diffuse glioma and whether MRI could be used to predict cognitive recovery postoperatively and during follow-up. Specifically, we wished to test the effect of tumours both locally and globally on the brain, in terms of their structural and functional network effects. We hypothesized that treatment-induced memory deficits are caused by disruption of brain networks that have been previously identified as fundamental for memory, and that this is mediated by global effects on normal brain microstructure.

## METHODS

### Sample

This study is a single centre prospective cohort design approved by the Cambridge Central Research Ethics Committee (protocol number 16/EE/0151). Patients deemed to have typical appearances of a diffuse glioma were identified at adult neuro-oncology multidisciplinary team (MDT) meetings at Addenbrooke’s Hospital (Cambridge, UK), and a consultant neurosurgeon directly involved in the study identified potential patients based on the outcome of the MDT discussion. All patients gave written informed consent. Inclusion criteria included: (*i*) participant is willing and able to give informed consent for participation in the study, (*ii*) imaging is evaluated by the MDT and judged to have typical appearances of a diffuse glioma, (*iii*) Stealth MRI is obtained (routine neuronavigation MRI scan performed prior to surgery), (*iv*) World Health Organisation (WHO) performance status 0 or 1, (*v)* age between 18 to 80 years, (*vi*) tumour located in or near eloquent areas of the brain thought to be important for speech and executive functions, and (*vii*) patient undergoing awake surgical resection of a diffuse glioma. This last inclusion criterion was adopted to collect additional intraoperative electrocorticography data that will be reported separately^21^. Participants were excluded if any of the following applied: (*i*) concomitant anti-cancer therapy, *(ii)* concomitant treatment with steroids, (*iii*) history of previous malignancy (except for adequately treated basal and squamous cell carcinoma or carcinoma in-situ of the skin) within 5 years, and (*iv*) previous severe head injury.

We recruited 17 patients aged 22-56 years (8 females). Final histological diagnoses revealed different grades of glioma: WHO-I n=2, WHO-II n=7, WHO-III n=5, WHO-IV n=3. Resection was complete (no residual FLAIR signal) in 9 patients, whereas 8 patients had partial resection. Adjuvant chemoradiotherapy was performed in 12 patients. Each patient was scanned up to four times: before surgery (preop), within 72 hours after surgery (postop), and at 3 and 12 months after surgery (month-3 and month-12). See **Table 1** for demographic details of participants.

**Table 1.**
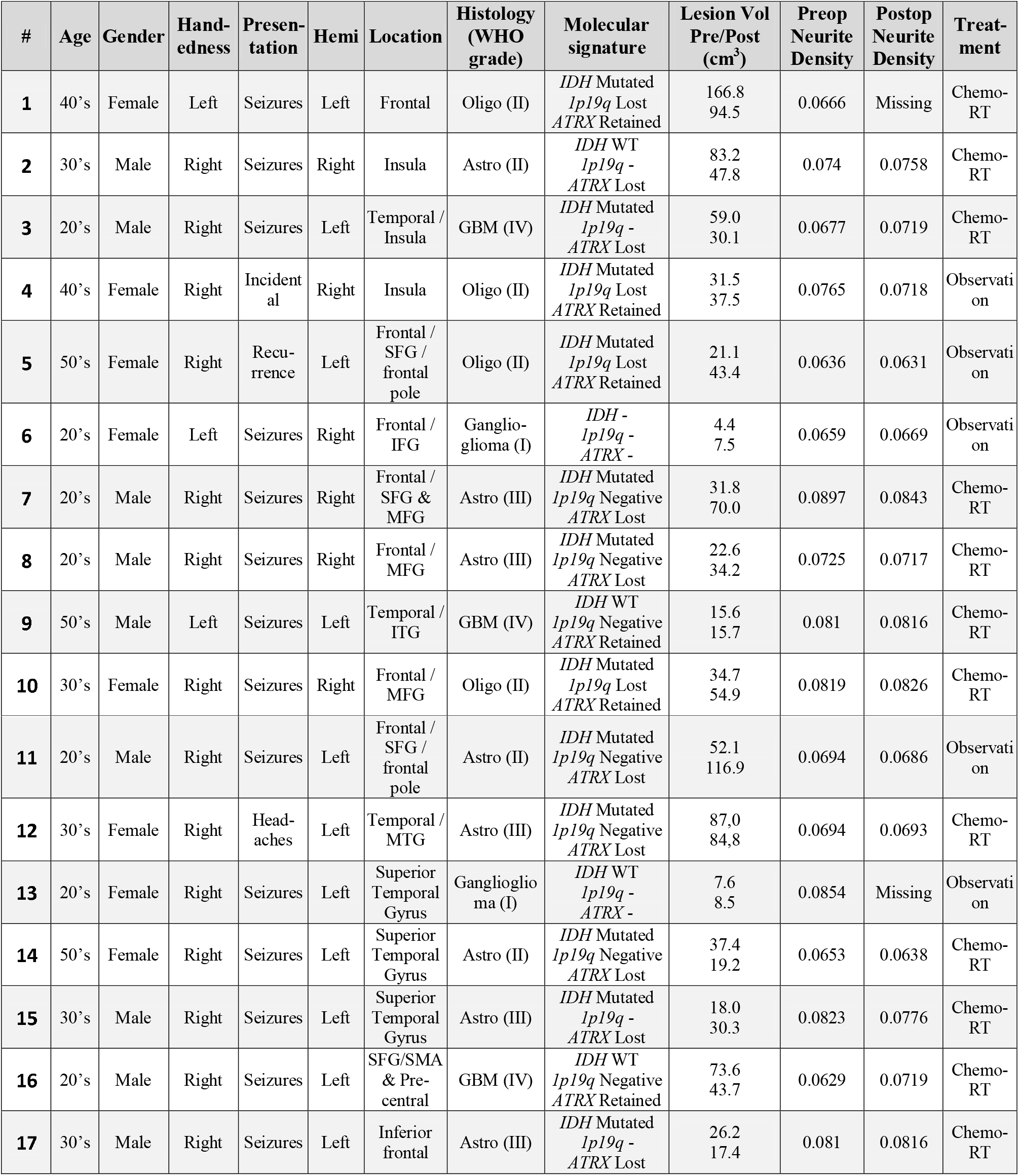
Demographic and pathological information. # Number of patient, SFG, Superior Frontal Gyrus; MFG, Middle Frontal Gyrus; IFG, Inferior Frontal Gyrus; ITG, Inferior Temporal Gyrus; MTG, Middle Temporal Gyrus; SMA, Supplementary Motor Area; RT, radiotherapy; Astro – astrocytoma; GBM – glioblastoma multiforme; Oligo – oligodendroglioma; Lesion Vol Pre/Post – Total volume occupied by the tumour (Pre) and total amount of damaged tissue (Post) according the lesion mask manually drawn on the MPRAGE image and refined with Unified Segmentation Lesion toolbox. Preop and Postop Neurite Density was calculated as the average Neurite Density for the whole brain of each individual.

| # | Age | Gender | Hand-<br>edness | Presen-<br>tation | Hemi | Location | Histology<br>(WHO<br>grade) | Molecular<br>signature | Lesion Vol<br>Pre/Post<br>(cm<sup>3</sup>) | Preop<br>Neurite<br>Density | Postop<br>Neurite<br>Density | Treat-<br>ment |
| --- | --- | --- | --- | --- | --- | --- | --- | --- | --- | --- | --- | --- |
| 1 | 40's | Female | Left | Seizures | Left | Frontal | Oligo (II) | IDH Mutated<br>1p19q Lost<br>ATRX Retained | 166.8<br>94.5 | 0.0666 | Missing | Chemo-RT |
| 2 | 30's | Male | Right | Seizures | Right | Insula | Astro (II) | IDH WT<br>1p19q -<br>ATRX Lost | 83.2<br>47.8 | 0.074 | 0.0758 | Chemo-RT |
| 3 | 20's | Male | Right | Seizures | Left | Temporal /<br>Insula | GBM (IV) | IDH Mutated<br>1p19q -<br>ATRX Lost | 59.0<br>30.1 | 0.0677 | 0.0719 | Chemo-RT |
| 4 | 40's | Female | Right | Incident-<br>al | Right | Insula | Oligo (II) | IDH Mutated<br>1p19q Lost<br>ATRX Retained | 31.5<br>37.5 | 0.0765 | 0.0718 | Observati-<br>on |
| 5 | 50's | Female | Right | Recur-<br>rence | Left | Frontal /<br>SFG /<br>frontal<br>pole | Oligo (II) | IDH Mutated<br>1p19q Lost<br>ATRX Retained | 21.1<br>43.4 | 0.0636 | 0.0631 | Observati-<br>on |
| 6 | 20's | Female | Left | Seizures | Right | Frontal /<br>IFG | Ganglio-<br>glioma (I) | IDH -<br>1p19q -<br>ATRX - | 4.4<br>7.5 | 0.0659 | 0.0669 | Observati-<br>on |
| 7 | 20's | Male | Right | Seizures | Right | Frontal /<br>SFG &<br>MFG | Astro (III) | IDH Mutated<br>1p19q Negative<br>ATRX Lost | 31.8<br>70.0 | 0.0897 | 0.0843 | Chemo-RT |
| 8 | 20's | Male | Right | Seizures | Right | Frontal /<br>MFG | Astro (III) | IDH Mutated<br>1p19q Negative<br>ATRX Lost | 22.6<br>34.2 | 0.0725 | 0.0717 | Chemo-RT |
| 9 | 50's | Male | Left | Seizures | Left | Temporal /<br>ITG | GBM (IV) | IDH WT<br>1p19q Negative<br>ATRX Retained | 15.6<br>15.7 | 0.081 | 0.0816 | Chemo-RT |
| 10 | 30's | Female | Right | Seizures | Right | Frontal /<br>MFG | Oligo (II) | IDH Mutated<br>1p19q Lost<br>ATRX Retained | 34.7<br>54.9 | 0.0819 | 0.0826 | Chemo-RT |
| 11 | 20's | Male | Right | Seizures | Left | Frontal /<br>SFG /<br>frontal<br>pole | Astro (II) | IDH Mutated<br>1p19q Negative<br>ATRX Lost | 52.1<br>116.9 | 0.0694 | 0.0686 | Observati-<br>on |
| 12 | 30's | Female | Right | Head-<br>aches | Left | Temporal /<br>MTG | Astro (III) | IDH Mutated<br>1p19q Negative<br>ATRX Lost | 87.0<br>84.8 | 0.0694 | 0.0693 | Chemo-RT |
| 13 | 20's | Female | Right | Seizures | Left | Superior<br>Temporal<br>Gyrus | Gangliogli-<br>oma (I) | IDH WT<br>1p19q -<br>ATRX - | 7.6<br>8.5 | 0.0854 | Missing | Observati-<br>on |
| 14 | 50's | Female | Right | Seizures | Left | Superior<br>Temporal<br>Gyrus | Astro (II) | IDH Mutated<br>1p19q Negative<br>ATRX Lost | 37.4<br>19.2 | 0.0653 | 0.0638 | Chemo-RT |
| 15 | 30's | Male | Right | Seizures | Left | Superior<br>Temporal<br>Gyrus | Astro (III) | IDH Mutated<br>1p19q -<br>ATRX Lost | 18.0<br>30.3 | 0.0823 | 0.0776 | Chemo-RT |
| 16 | 20's | Male | Right | Seizures | Left | SFG/SMA<br>& Pre-<br>central | GBM (IV) | IDH WT<br>1p19q Negative<br>ATRX Retained | 73.6<br>43.7 | 0.0629 | 0.0719 | Chemo-RT |
| 17 | 30's | Male | Right | Seizures | Left | Inferior<br>frontal | Astro (III) | IDH Mutated<br>1p19q -<br>ATRX Lost | 26.2<br>17.4 | 0.081 | 0.0816 | Chemo-RT |

### MRI and NODDI data acquisition and pre-processing

MRI data were acquired using a Siemens Magnetom Prisma-fit 3 Tesla MRI scanner and 16-channel receive-only head coil (Siemens AG, Erlangen, Germany). A T1-weighted MRI MPRAGE sequence was acquired using the following parameters: repetition time (TR) = 2300 ms, echo time (TE) = 2.98 ms, flip angle (FA) = 9 deg, 1 mm^3^ resolution, Field of View (FOV) = 256×240 mm^2^, 192 contiguous slices and acquisition time of 9 minutes and 14 seconds. During the same scanning session, we used a recently developed MRI multi-shell diffusion technique, NODDI with 30 directions (b-value=800 mm/s), 60 directions (b-value=2000 mm/s) and 10 unweighted B0 images. Other acquisition parameters were: TR = 8200 ms, TE = 95 ms, 2.5 mm^3^ voxel resolution, 60 slices, FOV = 240 mm and acquisition time of 15 minutes and 19 seconds. NODDI Matlab Toolbox (http://mig.cs.ucl.ac.uk/index.php?n=Tutorial.NODDImatlab) was used to quantify the microstructural complexity of dendrites and axons *in vivo*^11^. Compared with traditional DTI, this multi-compartment tissue model disentangles two key contributing factors of Fractional Anisotropy: the Gaussian contribution from water molecules located in the extracellular space (defined as the space around neurites), and the restricted non-Gaussian diffusion that takes place in the intra-cellular space that is bounded by axonal and dendritic membranes. The apparent intra-cellular volume fraction that represents the fraction of dendrites and axons was used here as a measurement of Neurite Density.

### Lesion masking and image transformation to standard space

Images were corrected for B0 field inhomogeneity, Gibbs artefacts and eddy-current distortions using MRtrix v3 (https://www.mrtrix.org/) and FSL v5.0 (http://fsl.fmrib.ox.ac.uk). Scans were visually inspected by an experienced researcher (RRG). Masks of the pre-op tumour and follow-up lesion (reflecting for example resected tissue, residual tumour, post-surgical oedema, or gliosis) masks were created using a semi-automated procedure. An experienced neurosurgeon (MH) initially did a manual delineation for each participant on the preoperative T1 image slices that included the tumour, and the resection site on the follow-up T1 images. Resulting masks were refined by the Unified Segmentation with Lesion toolbox (https://github.com/CyclotronResearchCentre/USwithLesion) that uses tissue probability maps to create a posterior tumour/lesion probability map. Inter-regional distances to the tumour boundary as defined in the tumour mask were estimated as the geodesic distance of the shortest path constrained by the white matter.

For each scan, the first B0 image of the diffusion sensitive sequence was linearly coregistered to the T1 image using ANTs (http://stnava.github.io/ANTs/). The resulting inverse transformation was used to map the Neurite Density map into the T1 image space. Each T1 image was non-linearly coregistered to standard space using Symmetric Normalization (ANTs-SyN), but excluding the tumour/lesion mask from the non-linear step of the wrapping to avoid distorting the spatial distribution of the tumour/lesion. The resulting transformation was additionally used to map the lesion mask and the Neurite Density map from T1 space to standard template space. The ICBM 2009a symmetric brain, an unbiased non-linear average of the MNI152, was used here as a standard template for normalization of Neurite Density using the contralateral values hemisphere that contained the tumour.

### Networks atlas based on normative data

We additionally utilised the map of large-scale networks defined in Yeo et al. (2011). This atlas was created by clustering functionally coupled regions in 1000 young, healthy adults. Regions delimited on the 7-Network liberal version of the Yeo atlas were used as Regions-of-Interest (ROIs) for calculating tumour overlap and Neurite Density (**Figure 1**, top). The visual network was not included in the analyses because none of the patients presented with a tumour in the occipital lobe. The limbic network was also excluded from further analyses due to its poor signal to noise ratio and lack of reproducibility^23^.

**Figure 1.**
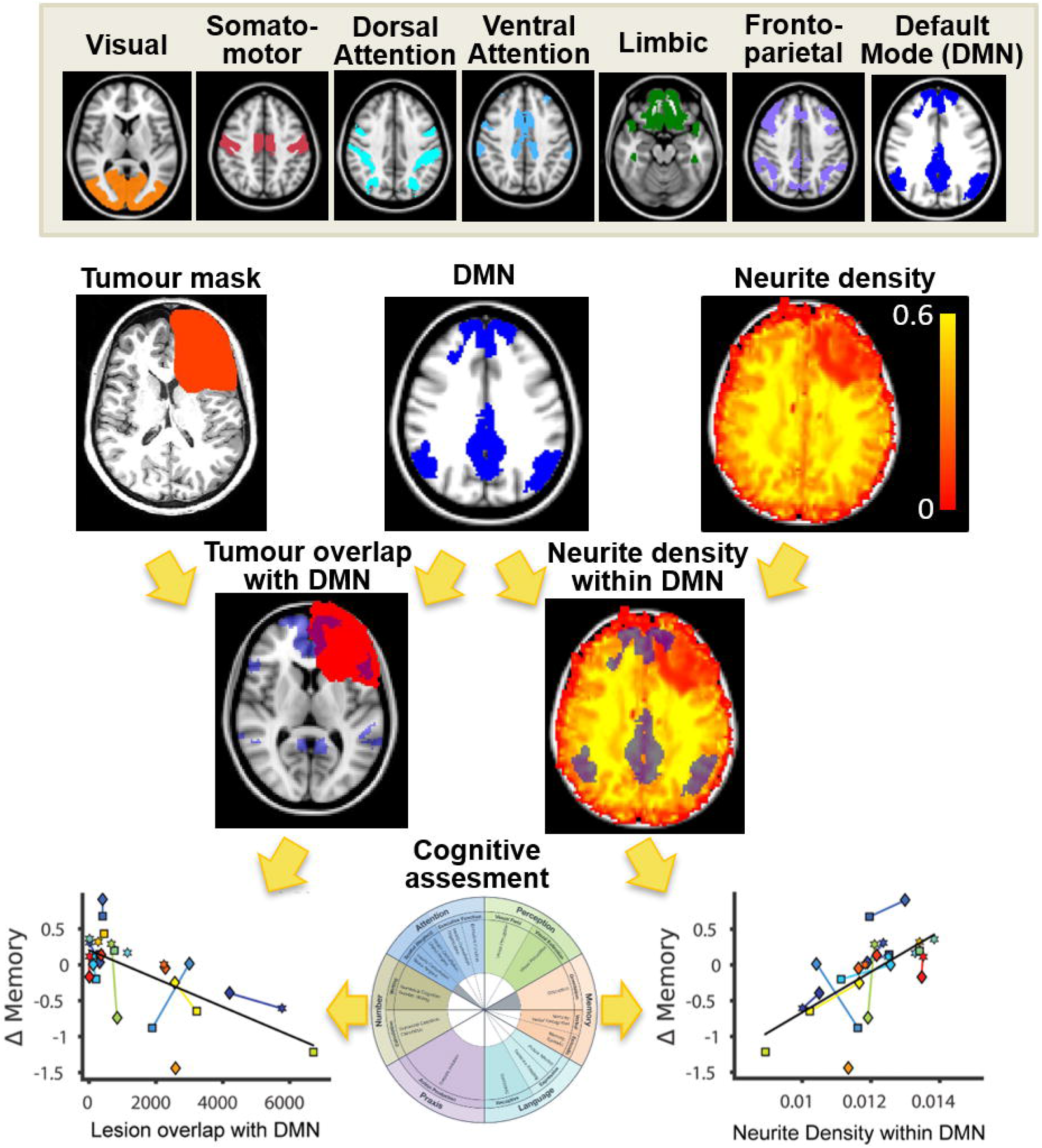
Flowchart of pipeline analysis. After transforming lesion masks to standard space, tumour/lesion spatial overlap and Neurite Density were calculated for each Yeo network (Yeo et al. 2011). For each assessment (preoperative, postoperative, 3 months and 12 months), lesion overlap and Neurite Density were compared with memory recovery. DMN – default mode network.

### Tumour overlapping and Neurite Density estimation

As location of the tumour varied across patients, we calculated, for each network and participant, the “Tumour Overlap” index as the amount of regional volume (in mm^3^) that spatially overlapped with the tumour (for preoperative assessments) or lesioned tissue (for postoperative and follow-up assessments) according to the tumour/lesion masks, after being transformed to standard space.

Median Neurite Densities in each Yeo network were calculated for each participant. Only voxels of the network that were not overlapping with the tumour/lesion masks were included in the analysis to reduce the impact of tumour volume on Neurite Density estimation. See **Figure 1** for an illustrative flowchart.

### Cognitive assessment

Immediately after each MRI scanning session, memory performance was evaluated using the Oxford Cognitive Screen (OCS)-bridge tablet-based a tool (https://ocs-bridge.com/), which is specifically designed for clinical setting. OCS-bridge is an app-based extension of the Birmingham Cognitive Screen and Oxford Cognitive Screen, two paper and pencil assessment tests that are widely used in health services in UK and that have been translated into multiple languages^24^.

We performed 6 memory screening tasks that included: free verbal memory, overall verbal memory, episodic memory, orientation, forward digit span and backwards digit span. For each individual task, a z-score value was computed by subtracting the mean and dividing by the standard deviation of task scores across participants. Tasks defined by an inverted scale where high values represent low memory performance were flipped by multiplying the z-score by -1. Total memory score was calculated as the average z-score of all tasks. By subtracting the preoperative z-score from the z-score of each subsequent task, we defined the longitudinal trajectory of each assessment (Δ) where negative scores represent worse performance than before surgery (*i*.*e*. memory deficit) and positive scores are associated with increased performance than before surgery (*i*.*e*. memory improvement). We have recently demonstrated that memory scores of these brain tumour patients are consistent between OCS-bridge and a traditional 2-3 hours neuropsychological interview^5^.

### Statistical analysis

Associations between cognition and imaging markers were tested using Linear Mixed Effect Models (LMMs) to account for the dependency of repeated measurements:

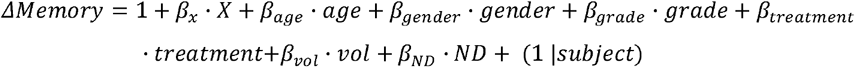

where *ΔMemory* represents the total memory score normalized to preoperative performance, *X* is the predictor variable based on imaging data (*i*.*e*. tumour overlap or Neurite Density for a given network) and (1 |*subject*) represents the random intercept of the model. The model included as covariates: age, gender, grade (low- vs. high-grade glioma), treatment (surgery alone vs. surgery plus chemo-radiotherapy), tumour volume and total Neurite Density (*ND*, only for predictions based on ND). All statistical tests were corrected for multiple comparisons using Benjamini–Hochberg False Discovery Rate (FDR < 0.05).

Missing data in limited sample size scenarios increases the chances of reporting false negatives. To partially address this issue, the main analyses were validated in an imputed dataset constructed from deriving missing imaging and cognitive assessments using a PCA-based tool for data inference (Missing Data Imputation toolbox, Trimmed score regression algorithm, 5000 iterations, tolerance < 10^−10^)^25^.

## RESULTS

### Memory recovery trajectories

Tumours were located on frontal (5 Left Hemisphere -LH- and 4 Right hemisphere -RH-), temporal (6 LH) and insular (2 RH) lobes (**Figure 2A, Table 1**). OCS-Bridge cognitive assessment was completed by 17 patients before surgery, 8 after surgery, 11 after 3 months and 11 after 12 months (47 in total). Seven patients could not tolerate postoperative screening and the remain missing assessments could not be collected due to logistic and technical reasons (14 out of 69). Years of education showed no correlation with preoperative memory performance (*R*^*2*^= 0.03, *P*=0.63) nor with memory recovery (*R*^*2*^= 0.02, *P*=0.50). Surgical resection and treatment had an impact on memory in most participants. Memory recovery after surgery (ΔMemory) showed a variety of trajectories, including progressive impairment, impairment followed by recovery, no change and improvement after surgery and during recovery.

**Figure 2.**
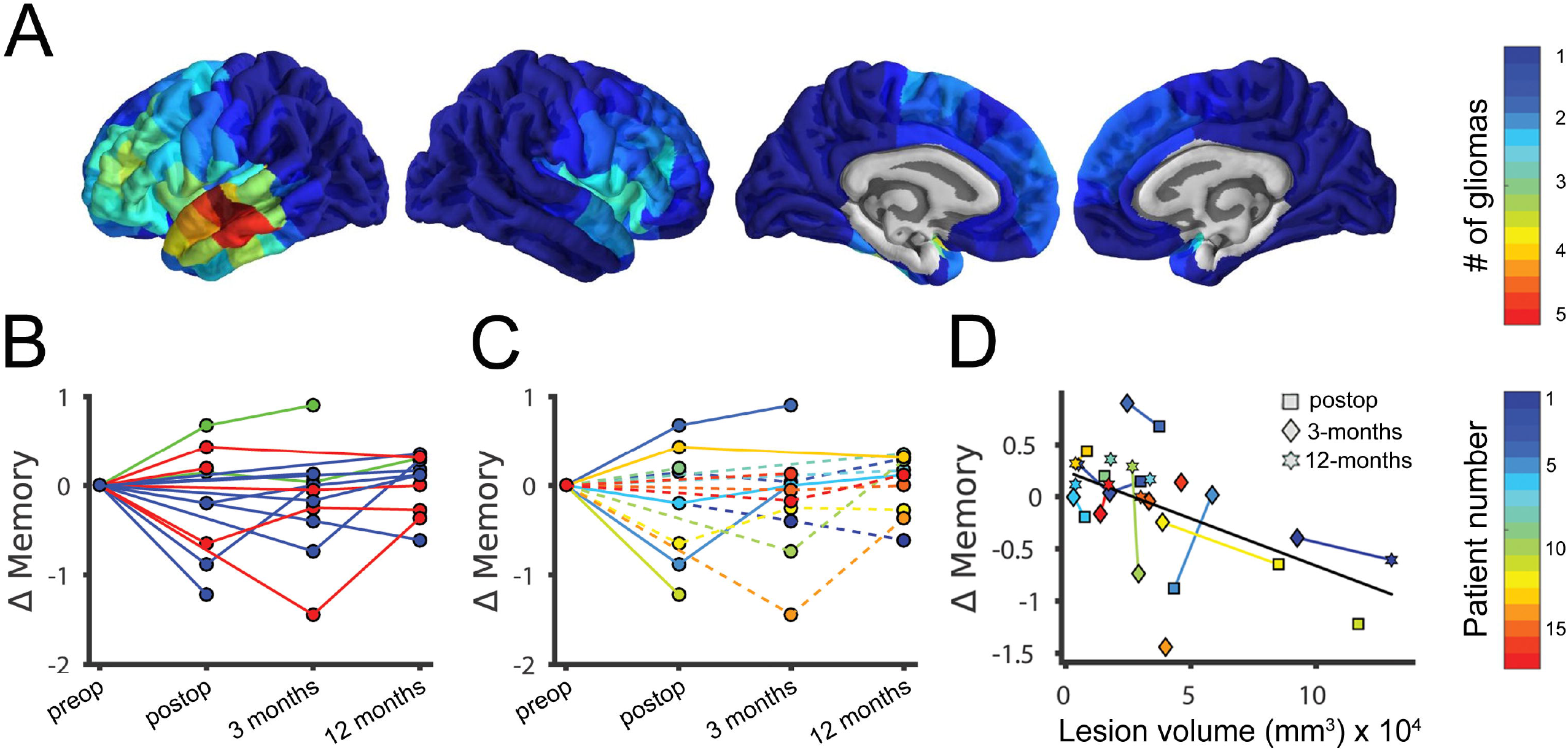
Patients’ memory recovery trajectories as a function of tumour location and type of treatment. **(a)** Tumour location in the cohort. Colour scale indicates the number of participants that had a tumour in a given cortical region. Cortical surface representations were plotted using BrainsForPublication v0.2.1 (https://doi.org/10.5281/zenodo.1069156). **(b)** Memory recovery trajectories normalised to pre-operative values coloured according to tumour location (frontal, blue; temporal, red; insular, green). **(c)** Recovery trajectories of patients receiving no further treatment beyond surgery (solid lines) and patients that also underwent chemo-radiotherapy (dashed lines). **(d)** Association between total lesion volume and memory recovery (postoperative, 3 months and 12 months) across patients. Memory recovery was normalized to pre-surgical values. Each color represents an individual participant as presented in **Table 1**. Positive scores correspond with post-surgical memory recovery and negative scores to post-surgical deficits.

### No impact of tumour location or volume on memory recovery

Follow-up recovery trajectories were not associated with tumour location based on traditional lobe classification (LMM, *F*_*val*_=0.05, *P=0*.*11*, **Figure 2B**) nor with the type of treatment received by patients (*i*.*e*. surgery alone or surgery with adjuvant chemoradiotherapy; LMM, *F*_*val*_<10^−3^, *P*=0.98, **Figure 2C**). The influence of the tumour and its treatment on recovery was initially evaluated by comparing the total lesion volume with changes in memory. Memory recovery was not correlated with tumour volume before surgery (*Spearmans’* ρ=-0.27, *P*=0.40), nor with lesion volume during recovery (LMM, *F*_*val*_=6.78, *P*=0.097, **Figure 2D**).

### Peri-tumoural Neurite Density within brain networks is associated with memory recovery

The impact of tumours and their treatment on brain structure were explored using a Neurite Density marker derived from diffusion imaging based on NODDI. Neurite Density showed no correlation with age (*R*^*2*^= 0.02, *P*=0.57), years of education (*R*^*2*^= 0.016, *P*=0.62), preoperative memory performance (*R*^*2*^= 0.002, *P*=0.87), nor with memory recovery (*R*^*2*^= 0.13, *P*=0.06), but was significantly different between networks in the hemisphere contralateral to the tumour (*F*_*val*_=42.56, *P*<10^−18^, **Figure S1**). We found that average whole-brain Neurite Density outside the tumour was negatively associated with tumour volume (*R*^*2*^= 0.28, *P*=0.029, **Figure 3A**), suggesting that tumours may mediate global effects on brain structural integrity. In support of this hypothesis, we found a distance-effect on Neurite Density as a function of distance to the tumour boundary. Peri-tumoural regions located between 0 and 20 mm outward from the tumour boundary had up to half of the Neurite Density compared to contralateral regions (**Figure 3B**), suggesting an extended tumour and brain structural integrity interactions. Preoperative Neurite Density within the DMN in both hemispheres (*i*.*e*. not normalized) was positively correlated with memory performance during recovery, suggesting that Neurite Density has an initial protective effect on outcome (*F*_*val*_=13.2, *P*_*fdr*_=0.001, **Figure 3C**). Moreover, Neurite Density during recovery was also associated with memory performance. When postoperative and follow-up Neurite Density values were considered, Memory scores were correlated with Neurite Density within fronto-parietal (*F*_*val*_=9.62, *P*_*fdr*_=0.01) and DMN (*F*_*val*_=21.5, *P*_*fdr*_<10^−3^) (**Figure 3D**). Neither preoperative nor follow-up Neurite Density with any of the remain networks were associated with memory recovery (all *P*_*fdr*_>0.05; **Table S1**). All associations were tested using LMM after regressing out age, gender, tumour grade, total lesion volume, type of treatment and total Neurite Density effects. Results shown in Figure 3C and 3D were validated using an imputed dataset where missing assessments were inferred from the available data (**Figure S2**).

**Figure 3.**
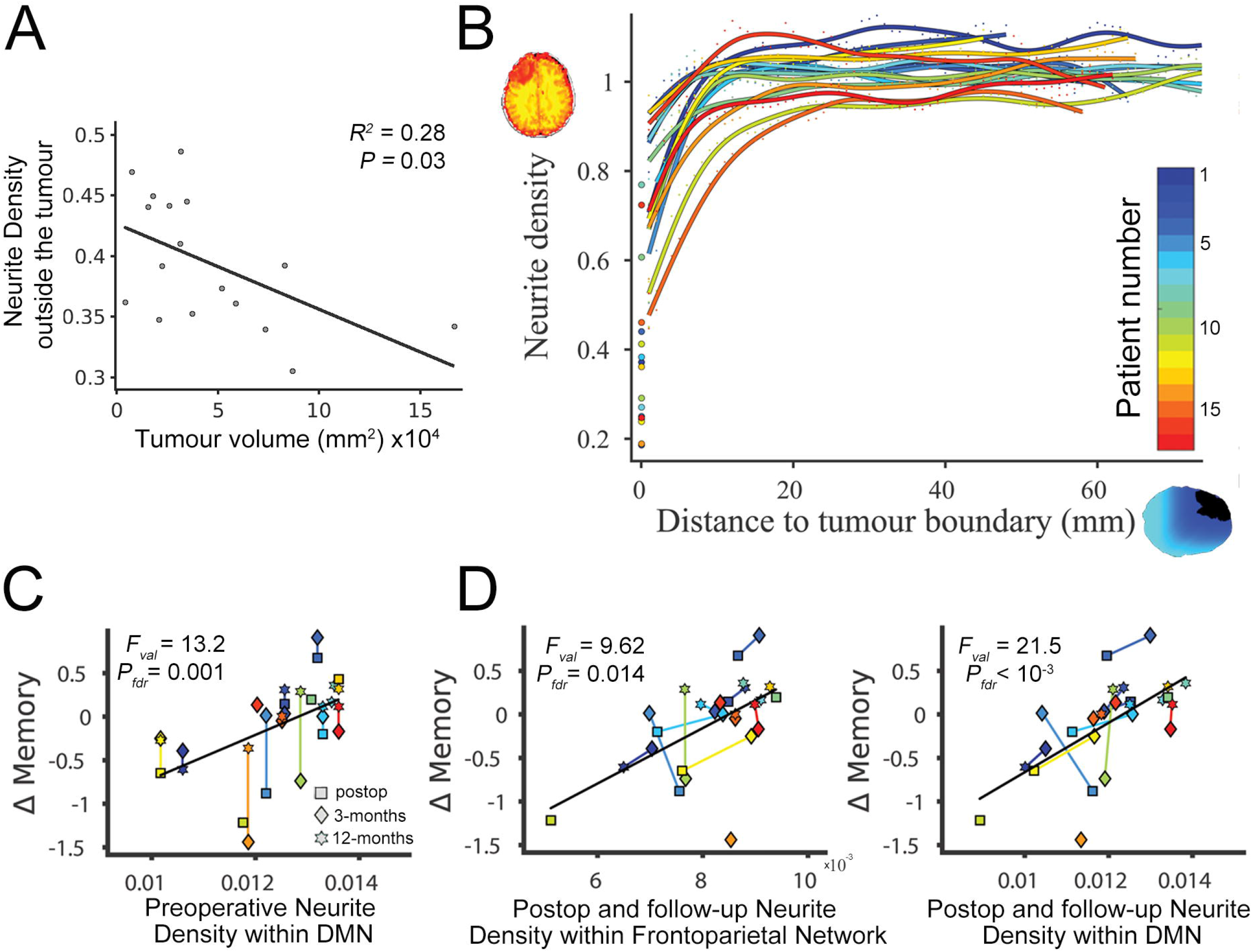
Impact of tumour and tumour treatment in Neurite Density and memory recovery. **(a)** Association between total tumour volume and Neurite Density outside of the tumour. **(b)** Neurite Density as a function of distance to the tumour boundary. Values are normalized to the contralateral hemisphere (*i*.*e* values less than one represent regions with reduced neurite density). Zero distance (x=0 mm) corresponds to average Neurite Density values within each tumour. Each color represents an individual participant. Due to high-dimensional data, error bars are too small to be displayed. **(c)** Memory recovery (postoperative, 3 months and 12 months) as a function of *preoperative* Neurite Density within the DMN. For a given participant, the same Neurite Density (preoperative) value was used in all assessments, resulting in vertical lines between them. **(d)** Memory recovery as a function of *postoperartive and follow-up* Neurite Density within the fronto-parietal and DMN. Neurite Density corresponds with the median density within the White Matter of each network, excluding lesioned regions. Colors illustrate each participant with lines connecting them.

### Tumour overlap with the DMN is associated with memory recovery

Preoperative tumour overlap with the DMN was negatively correlated with memory recovery (*F*_*val*_=12.7, *P*_*fdr*_=0.002; **Figure 4A**). When follow-up imaging data were included in the analyses (post-operative, 3 months and 12 months), we also found a significant association between lesion overlapping with the Default Mode Network (DMN) and memory performance (*F*_*val*_=31.6, *P*_*fdr*_<10; **Figure 4B**). Neither (preoperative) tumour nor (follow-up) lesion overlapping with any of the remain networks were associated with memory recovery (all *P*_*fdr*_>0.05; **Table S1**). All associations were tested using LMM after regressing out age, gender, tumour grade, type of treatment and total lesion volume effects. Results were validated using an imputation for missing data (**Figure S2**).

**Figure 4.**
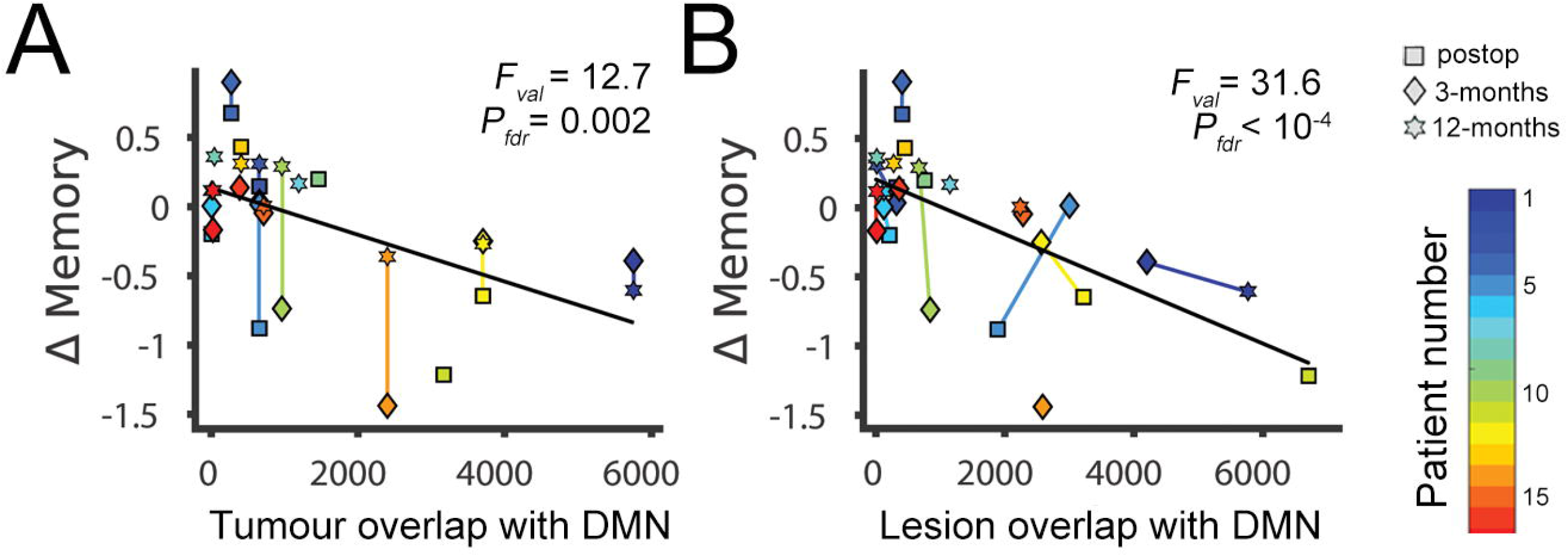
Memory recovery as a function of lesion overlapping with DMN. **(a)** Memory recovery as a function of tumour overlap with DMN. For a given participant, the (preoperative) tumour overlap value was used in all assessments, resulting in vertical lines between them. **(b)** Memory recovery a function of (follow-up) lesion overlap with DMN. Overlap values were defined as the number of voxels (equivalent to mm^3^) of the tumour mask that overlapped with each network. Lines link assessments of the same participant.

## DISCUSSION

In this study, we combined structural and NODDI diffusion MRI with normative brain network data from healthy participants and whole-brain analyses to determine whether memory recovery trajectories are affected by the impact that the tumour and its treatment have on specific circuits. Overall, our results suggest that the effect of a tumour on the brain and its consequences in memory depends on interactions with the DMN at both global levels. Acquiring high-quality longitudinal memory outcome data required nuanced study design. Using a tablet-based cognitive assessment app afforded specific advantages, such as avoiding reliance on having a trained neuropsychologist available (with associated time and financial costs), more easily facilitating follow-up screening, and collecting specific data that is difficult to acquire using traditional clinical interviews tests, such as accurate reaction times and interactive visuospatial paradigms. We observed a variety of memory recovery trajectories after surgery and during subsequent treatment that included: no change, postsurgical deterioration and then improvement, and postsurgical improvement and then deterioration. Although the mechanisms behind cognitive improvement after major surgery are not understood, these findings are not without precedent^26^. Nevertheless, given that pre- and postoperative assessments were performed in a relatively short period of time, we cannot discard the possibility of practice and learning effects on the tasks.

Tumour location is one of the most relevant features to be considered when estimating the cognitive risks of surgical resection. In our cohort, gliomas were mainly located in frontal, temporal and insular cortices on the left hemisphere. The higher prevalence of left-hemisphere tumours is because this study only included patients undergoing awake brain surgery. On the other hand, the higher prevalence of frontal, temporal and insular tumours is consistent with previous studies showing that low- and high-grade gliomas are relatively scarce in primary cortices and occipital lobes^27^. Although several developmental, cytomyeloarchitectonic, neurochemical, metabolic, and functional reasons have been proposed, the mechanisms behind the preferential location of gliomas in frontal, temporal and insular cortices is still an ongoing debate^28^. The presence of gliomas in secondary and association cortices that have been traditionally associated with cognitive processing^29^ may be an important factor to understand cognitive deficits induced by the tumour and its treatment.

Beyond the impact that age, gender, tumour grade, tumour volume and type of treatment have on recovery, we hypothesized that memory functioning was associated with treatment-related interactions with brain networks that have been previously identified as fundamental for cognition and memory. In support of this hypothesis, we found significant associations between memory recovery and lesion overlap with the DMN. The DMN constitutes a set of regions that are active during passive rest and it is considered the neuronal correlate of internally-generated self-referential cognition. Accordingly, a large body of literature has accumulated evidence about the role of the DMN in memory functioning, including semantic^30^, episodic^31^, autobiographical^32^ and working memory^33^. Damage to the DMN has been associated with memory deficits in patients with brain lesions^32^ and traumatic brain injury^34^. To the best of our knowledge, there are no studies linking DMN disruption with memory recovery in brain tumour patients to date, but decreases of functional connectivity in this network have been extensively reported in the literature. DMN functional connectivity is reduced in glioma patients when compared with controls in both the hemisphere ipsilateral and contralateral to the tumour, which is particularly prominent for tumours located on the left side of the brain^35^.

Neurite Density derived from NODDI has been previously associated with age^36,37^ and gender^38^. However, to the best of our knowledge there is no study formally exploring associations between Neurite Density and years of education or education level. Not surprisingly, we found a consistent Neurite Density decrease within the tumour. Given that Neurite Density derived from NODDI and Fractional Anisotropy are strongly correlated^11^, our results align with previous evidence found from Diffusion Tensor Imaging (DTI) studies showing that glioblastomas have reduced Fractional Anisotropy compared with the rest of the brain^39^. For its part, NODDI has shown higher sensitivity for glioma grade differentiation than other diffusion sequences^40^. Zhao et al. (2018) recently reported that NODDI in combination with patient age can predict glioma grade with a sensitivity and specificity of 92% and 89%, respectively^18^. Despite these promising findings, the potential of NODDI as a predictor of patient cognitive recovery has not yet been explored in the literature.

Despite peri-tumoural regions being identified on T1 images as unaffected by the experienced neurosurgeon that delineated the mask, and by the semi-automatic segmentation procedure, these regions had decreased Neurite Density compared with the contralateral hemisphere. Long-distance tumour effects have been observed with regard to functional connectivity and functional complexity^41^. Disrupted white matter integrity has been reported in DTI studies that show decreased FA in peri-tumoural regions^42^ and white matter tracts^43^. Extratumoural pre- and post-operative Neurite Density within the fronto-parietal and DMN had a protective factor in memory recovery that may be mediated by the prominent role of these networks in this cognitive domain. From the connectomic perspective, cognitive performance of brain tumour patients has been associated with hub-related structural connectivity within the DMN and fronto-parietal Network in hemispheres contralateral to the tumour^44^. By using connectomic metrics derived from both DTI and fMRI, Liu et al. (2016) achieved a 75% accuracy when predicting survival of high-grade glioma patients^45^. However, further research is needed to better understand the mechanisms by which low- and high-grade glioma, and treatment disruption mediate memory and, more generally, cognitive recovery.

### Limitations

Brain tumours have a unique blood-oxygen-level-dependent (BOLD) fMRI signal compared with normal brain tissue^46,47^. For this reason, correlation-based time-series analysis, such as seed-based or Independent Component Analysis (ICA), are problematic unless tumoural tissue is excluded from the analysis. In the present study, including tumoural regions would have impeded the estimation of its overlap with each resting-state networks. Consequently, brain networks were defined using normative data from healthy individuals^22^. However, by using brain network templates in standard space, neglect the potential spatial shift of brain functioning. Thus, our results should be interpreted as the potential of the tumour to disrupt the corresponding healthy network, not the actual tumour/lesion overlap with each participant’s network.

LMMs can accommodate missing data, prevent false-positive associations due to population structure, and increase power by applying a structure-specific correction. However, 17 patients still represent a limited sample size for a heterogeneous condition. For this reason, it is possible that some of the reported non-significant associations were Type II errors. Moreover, treatment heterogeneity across patients also limits the generalisability of the results. As treatment was decided solely on clinical criteria, 6 patients had only a surgical intervention while 11 additionally had different chemo-radiotherapy regimes. Chemo-radiotherapy has a dose-dependent effect on white matter structural integrity that has been associated with poor cognitive performance^48^. Additionally, all our patients had the pre-operative imaging appearances of a diffuse glioma (for example, non-enhancing and without oedema or mass effect), subsequent pathological examination revealed a range of histological diagnoses, in keeping with previous studies demonstrating the diagnostic limitations of standard MRI sequences. Although the LMMs considered here regressed out the effect of age, gender, tumour grade, tumour volume and treatment, the individual contribution of these factors cannot be untangled here due to the limited sample size.

## Conclusion

Our findings highlight that memory recovery is not associated with the type of treatment, tumour volume nor the tumour location based on traditional lobes. Interestingly, Neurite Density is decreased beyond the tumour boundary and high values within fronto-parietal and DMN are associated with better memory recovery. On the other hand, tumour and lesion overlapping with DMN have a negative impact on memory. Taken together, these results reveal the potential of combining brain network analyses, normative connectomes, and advanced MRI sequences to better understand and predict the impact of brain tumours and their treatment on patients’ cognitive outcomes.

## Data Availability

Data is not currently available

## ACKNOWLEDGEMENTS

We thank all patients for generous involvement in the study. We also thank to Luca Villa, Jessica Ingham, Alexa Mcdonald for their contribution to the study. This research was supported by The Brain Tumour Charity and the Guarantors of Brain.

## DATA AVAILABILITY STATEMENT

In accordance with ethics requirements, data will be made available to collaborating centres upon reasonable request.

