## Supplemental Information for "Memory recovery is related to default mode network impairment and neurite density during brain tumours treatment"

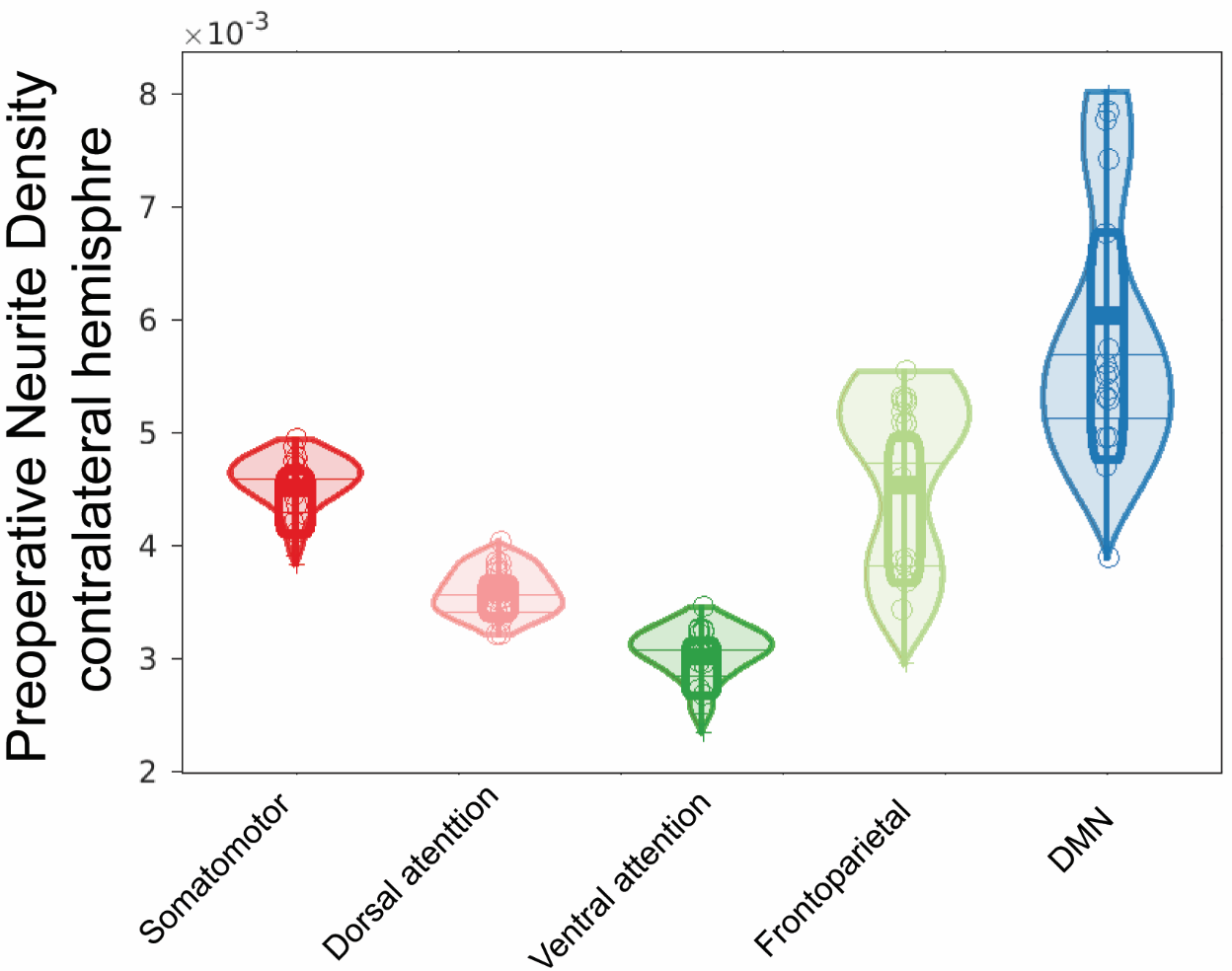

**Figure S1. Distribution of preoperative Neurite Density in the hemisphere contralateral to the tumour across the five Yeo networks considered.** *Post hoc t-tests* revealed a significant difference between each pair of networks (all *P*<10^-4^, FDR corrected), except between somatomotor and fronto-parietal (*P*=0.87).

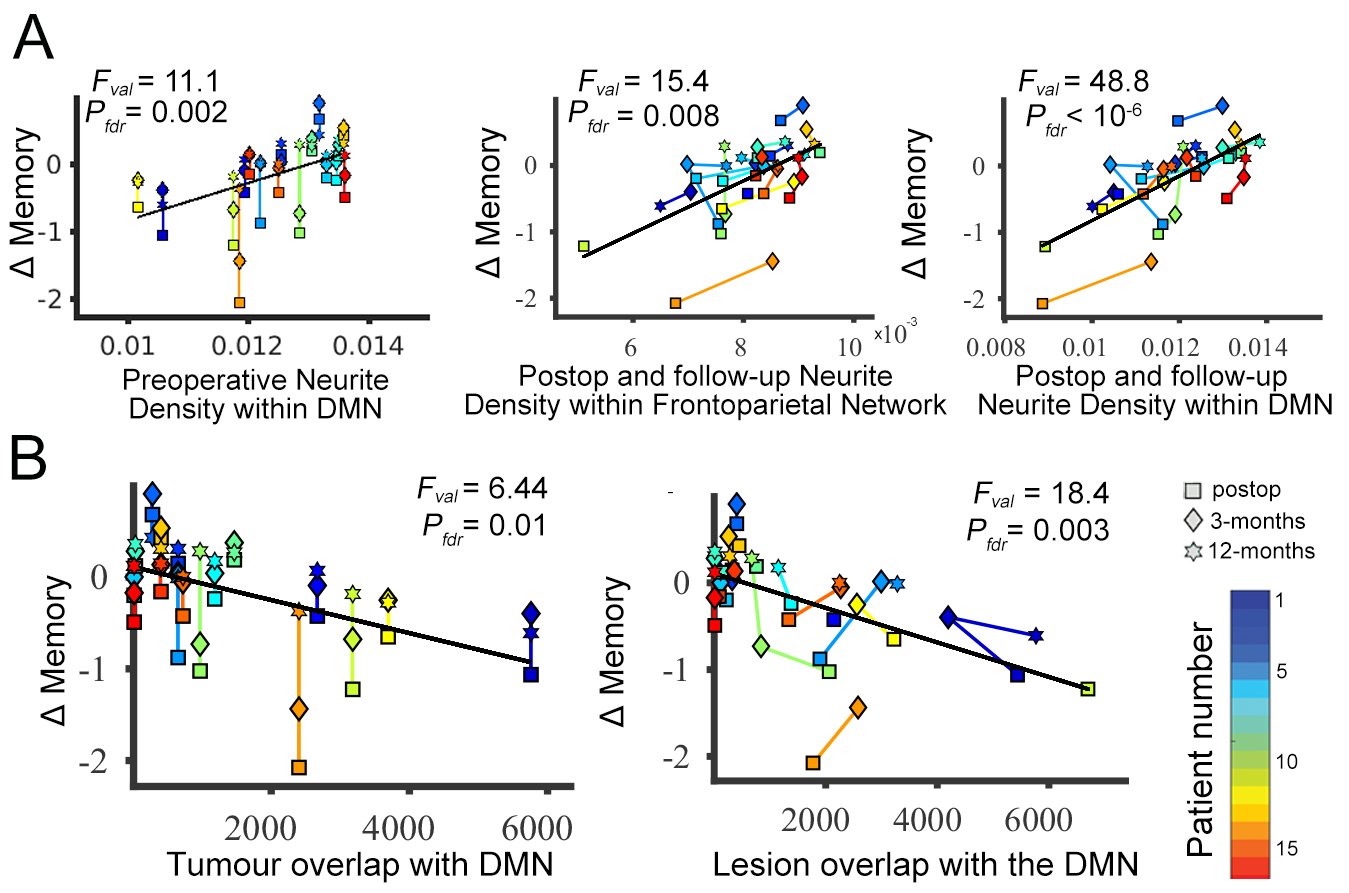

**Figure S2. Memory recovery as a function of Neurite Density and lesion overlapping with the DMN and fronto-parietal network using Multiple Imputation of missing data. (a)** Memory recovery as a function of preoperative (left) and postoperative and follow-up Neurite Density within the fronto-parietal Network (middle) and DMN (right), excluding lesioned regions **(b)**. Memory recovery as a function of preoperative (left) and postoperative and follow-up (right) lesion overlap with DMN. LMM calculated using age, gender, tumour grade, tumour volume and treatment. Missing data were imputed using the Missing Data Imputation (MDI) toolbox^1^. Missing data points (*i.e.* less than three assessments per patient) correspond with memory or imaging values that did not converge below the tolerance threshold of the imputation method. None of the other networks showed any significant association that survived correction for multiple comparisons.

|  | **Preop Neurite Density**  (Figure 3C) | | **Follow-up Neurite Density**  (Figure 3D) | | **Preop tumour overlapping**  (Figure 4A) | | **Follow-up tumour overlapping**  (Figure 4B) | |
| --- | --- | --- | --- | --- | --- | --- | --- | --- |
|  | ***F_value_*** | ***P_fdr_*** | ***F_value_*** | ***P_fdr_*** | ***F_value_*** | ***P_fdr_*** | ***F_value_*** | ***P_fdr_*** |
| **Somatomotor** | 0.56 | 0.47 | 0.75 | 0.47 | 0.02 | 0.89 | 3.38 | 0.16 |
| **Dorsal Attention** | 0.53 | 0.47 | 0.55 | 0.47 | 0.43 | 0.65 | 0.04 | 0.84 |
| **Ventral Attention** | 1.73 | 0.34 | 1.45 | 0.41 | 4.26 | 0.13 | 0.18 | 0.84 |
| **Fronto-parietal** | 3.33 | 0.20 | **9.62** | **0.01** | 0.61 | 0.65 | 2.97 | 0.16 |
| **DMN** | **13.3** | **<10^-3^** | **21.5** | **<10^-3^** | **12.7** | **<10^-3^** | **31.6** | **<10^-4^** |

**Table S1. Effect sizes and corrected *P-values* for all imaging-memory associations explored in the main manuscript.**
